# Plasma biomarkers for diagnosis of Alzheimer’s disease and prediction of cognitive decline in individuals with mild cognitive impairment

**DOI:** 10.1101/2022.04.18.22272912

**Authors:** Pia Kivisäkk, Thadryan Sweeney, Becky C. Carlyle, Bianca A. Trombetta, Kathryn LaCasse, Leena El-Mufti, Idil Tuncali, Lori B. Chibnik, Sudeshna Das, Clemens R. Scherzer, Keith A. Johnson, Bradford C. Dickerson, Teresa Gomez-Isla, Deborah Blacker, Derek H. Oakley, Matthew P. Frosch, Bradley T. Hyman, Anahit Aghvanyan, Pradeepthi Bathala, Christopher Campbell, George Sigal, Martin Stengelin, Steven E. Arnold

**Affiliations:** Alzheimer’s Clinical and Translational Research Unit, Department of Neurology, Massachusetts General Hospital, Harvard Medical School, Boston, MA; Precision Neurology Program and Center for Advanced Parkinson Research, Harvard Medical School, Brigham and Women’s Hospital, Boston, MA; Department of Neurology, Massachusetts General Hospital, Harvard Medical School, Boston, MA; Department of Epidemiology, Harvard TH Chan School of Public Health, Boston, MA; Department of Psychiatry, Massachusetts General Hospital, Harvard Medical School, Boston, MA; Department of Pathology, Massachusetts General Hospital, Harvard Medical School, Boston, MA; Meso Scale Diagnostics, LLC., Rockville, MD

## Abstract

**Background:** The last few years have seen major advances in blood biomarkers for Alzheimer’s Disease (AD) with the development of ultrasensitive immunoassays, promising to transform how we diagnose, prognose, and track progression of neurodegenerative dementias.

**Methods:** We evaluated a panel of four novel ultrasensitive electrochemiluminescence (ECL) immunoassays against presumed CNS derived proteins of interest in AD in plasma [phosphorylated-Tau181 (pTau181), total Tau (tTau), neurofilament light (NfL), and glial fibrillary acidic protein (GFAP)]. 366 plasma samples from the Massachusetts Alzheimer’s Disease Research Center’s longitudinal cohort study were examined to differentiate definite AD, other neurodegenerative diseases (OND), and cognitively normal (CN) individuals. A subset of samples were selected to have longitudinal follow up to also determine the utility of this plasma biomarker panel in predicting 4-year risk for cognitive decline in individuals with different levels of cognitive impairment.

**Results:** pTau181, tTau and GFAP were higher in AD compared to CN and OND, while NfL was elevated in AD and further increased in OND. pTau181 performed the best (AD vs CN: AUC=0.88, 2-fold increase; AD vs OND: AUC=0.78, 1.5-fold increase) but tTau also showed excellent discrimination (AD vs CN: AUC=0.79, 1.5-fold increase; AD vs OND: AUC=0.72, 1.3-fold increase). Participants with MCI who progressed to AD dementia had higher baseline plasma concentrations of pTau181, NfL, and GFAP compared to non-progressors with the best discrimination for pTau181 (AUC=0.82, 1.7-fold increase) and GFAP (AUC=0.81, 1.6-fold increase).

**Conclusions:** These new ultrasensitive ECL plasma assays for pTau181, tTau, NfL, and GFAP detect CNS disease with high specificity and accuracy. Moreover, the absolute baseline plasma levels of pTau and GFAP reflect clinical disease aggressiveness over the next 4 years, providing diagnostic and prognostic information that may have utility in both clinical and clinical trial populations.

**Classification of Evidence:** This study provides Class II evidence that plasma levels of pTau181, tTau, NfL, and GFAP are associated with AD and that pTau181 and GFAP are associated with progression from MCI to AD dementia.

## BACKGROUND

The recent emergence of ultrasensitive immunoassays for measuring biomarkers for Alzheimer’s Disease (AD) in blood holds promise to transform how we diagnose, prognose and track progression of AD. AD is a challenging disease to diagnose based on clinical presentation only^1^, and while CSF- and PET-based biomarkers are highly predictive of the presence of AD pathology in the brain^2^ and considered supportive evidence in recent diagnostic criteria^3^, their perceived invasiveness, high cost, and/or lack of broad availability renders them less suitable for the large number of individuals with cognitive complaints who are encountered in primary care and other non-specialized settings. Recent assay development has resulted in assays sensitive enough to reliably measure the classic amyloid-β (A), tau (T), and neurodegeneration (N) biomarkers, which provide the foundation of the current National Institute on Aging and Alzheimer’s Association (NIA-AA) research framework for diagnosing AD^3^, not only in CSF but also in blood^4^, circumventing many of the limitations of CSF and PET biomarkers.

While much progress has been made, there is a need to develop and optimize additional plasma biomarker assays on different platforms and to understand their performance and utility for clinical research, and ultimately, clinical practice. In this paper, we evaluated a panel of four novel ultrasensitive electrochemiluminescence (ECL) immunoassays from Meso Scale Diagnostics^®^ (MSD; Rockville, MD) of interest in AD: phosphorylated-Tau181 (pTau181), total Tau (tTau), neurofilament light (NfL), and glial fibrillary acidic protein (GFAP). Various pTau isoforms, such as pTau181, pTau217, and pTau231, appear thus far to be among the most promising AD biomarkers in plasma. These have repeatedly shown a diagnostic accuracy for the separation of AD from non-AD dementia (as classified by CSF or PET) with AUCs in some instances well over 0.85^5-9^. They have also shown good ability to predict progression from mild cognitive impairment (MCI) to AD dementia in individual patients^8, 10^. NfL, considered a general marker for neuroaxonal injury, is increased in plasma not only in AD but also in a range of other neurodegenerative disorders, stroke, head trauma, and other diseases^11^. Studies in multiple sclerosis and spinal muscular atrophy have shown that decreases in plasma NfL correlated with clinical improvement in treatment trials with different disease modifying therapies^12, 13^ providing proof of concept that plasma NfL can be used as surrogate marker for neurodegeneration in clinical trials. GFAP is an intermediate filament found in the cytoskeleton of astrocytes and is commonly used as a neuropathology marker for astrocyte activation in response to brain injury, neuroinflammation, and other CNS stress^14^. Recent studies have shown that plasma GFAP is elevated early in AD and continues to increase as the disease progresses^15, 16^, but its utility as a biomarker for AD is not yet fully established.

We used banked plasma samples from participants in the Massachusetts Alzheimer’s Disease Research Center’s longitudinal cohort (MADRC-LC) and evaluated the performance of pTau181, tTau, NfL, and GFAP measured using the novel MSD ECL assays. We first evaluated their ability to differentiate individuals with an autopsy confirmed, amyloid PET, and/or CSF AD biomarker-based diagnosis of AD from non-AD neurodegenerative diseases and cognitively normal individuals (CN). We also evaluated their usefulness in predicting cognitive decline in older individuals with no cognitive impairment, subjective cognitive impairment (SCI) or MCI.

## MATERIALS AND METHODS

### Study population

We included a total of 366 participants in the MADRC-LC study, a longitudinal observational study of cognitive aging, AD and AD-related disorders. Annual assessments include a general and neurological exam, a semi-structured interview with the participant and/or informant to record cognitive symptoms and score the Clinical Dementia Rating scale (CDR Dementia Staging Instrument), a battery of neuropsychological tests^17, 18^, and blood collection for all consenting participants. Cognitive status and clinical diagnosis are determined at each visit by a consensus team after a detailed examination and review of all available information according to 2011 NIA-AA diagnostic criteria for MCI^19^ and AD^20^. A subset of participants undergoes imaging and/or CSF biomarker substudies in affiliated protocols and all participants are invited to join a brain donation program.

Two partially overlapping samples were studied:

1. Sample A was assembled as a “high-contrast” diagnostic sample consisting of: a) 95 AD patients with autopsy and/or molecular biomarker-confirmation. Of these, 75 had intermediate or high AD neuropathologic changes upon autopsy according to the NIA-AA guidelines for the neuropathologic assessment of Alzheimer’s disease^21^, 14 had positive [11C]Pittsburgh Compound-B amyloid PET imaging, and 6 had positive CSF biomarkers [decreased amyloid ß42/40 ratio and increased total Tau and pTau181]^3^. The time interval was 4.0±2.4 years between plasma collection and death for autopsied individuals, 0.8±0.8 years between plasma collection and PET imaging, and 2.4±2.8 years between plasma and CSF collection; b) 53 OND participants with a variety of other neurodegenerative diseases and minimal to no AD neuropathological changes on autopsy. Their autopsy diagnoses were Frontotemporal Lobar Degeneration (FTLD) TDP43 (n=13), FTLD tau [progressive supranuclear palsy (n=10), Pick’s disease (n=6), corticobasal degeneration (n=3)], Lewy body disease (n=7), cerebrovascular disease (n=5), amyotrophic lateral sclerosis (n=4), Creutzfeldt-Jakob disease (n=1), cerebral amyloid angiopathy (n=1), multiple sclerosis (n=1), thalamic degeneration (n=1), and dementia lacking distinctive histology (n=1). The time between plasma collection and death was 2.8±1.9 years; and c) 90 cognitively normal controls (CN) with normal neuropsychological testing scores and no subjective cognitive symptoms during 8.8±3.7 years of follow-up.
2. Sample B was assembled as a longitudinal prognostic sample defined by cognitive trajectories over at least five annual follow-up visits over 4 years. 85 participants had a baseline clinical diagnosis of MCI due to probable AD and a global CDR score of 0.5 and were subclassified into two groups based on their CDR trajectory: MCI-decline (n=47) if their global CDR score increased from 0.5 to ≥1 during follow-up, and MCI-stable (n=38) if there was no change in global CDR score. 49 participants had a baseline clinical status of SCI and were classified into two groups based on their cognitive status trajectory: SCI-decline (n=19) if they were diagnosed as MCI for at least two follow-up visits or as dementia for at least one or more follow-up visits (and remained MCI or dementia at the most recent visit), and SCI-stable (n=30) if their cognitive status was SCI at the last follow-up visit and they had no more than one visit with a transient status of MCI. 53 participants had no cognitive complaints and a global CDR score of 0 at baseline and were classified into two groups based on their global CDR trajectory: CN-decline (n=14) if their global CDR score increased from 0 to 0.5 and they received a clinical diagnosis of MCI due to probable AD for at least two follow-up visits, and CN-stable (n=39) if they never had a global CDR score >0. Three CU-decline individuals progressed further from MCI to AD dementia with a global CDR score of 1 in the next few years following the end of the 4-year study window. 19 participants in the longitudinal sample had autopsy or CSF data (14 MCI-decline, 2 MCI-stable, 2 CU-decline, and 1 SCI-stable), all of which confirmed a diagnosis of AD.

### Standard Protocol Approvals, Registrations, and Patient Consents

The study was approved by the Mass General Brigham Institutional Review Board (2006P002104) and all participants or their assigned surrogate decision makers provided written informed consent.

### Plasma sampling and analysis

Banked plasma samples collected between 2008-2019 were obtained from the Harvard Biomarkers Study Biobank^22^. Samples were collected in K2EDTA tubes, centrifuged at 2000g for 5 min, frozen in low retention polypropylene cryovials within 4 hours of collection, and stored at −80°C until use. Ultrasensitive MSD S-PLEX^®^ assay kits (Meso Scale Discovery, Rockville, MD) employing a sandwich immunoassay format using monoclonal antibodies and ECL detection were used to detect plasma biomarker levels. pTau181 was measured using a commercial assay (catalog # K151AGMS) following manufacturer’s instructions while prototype S-PLEX assays were used for NfL, GFAP and tTau. The kits used MSD’s ultrasensitive S-PLEX ECL format, which provided additional signal enhancement and sensitivity relative to conventional ECL formats^23^. Calibrators for the different assays were prepared by using recombinant Tau441 expressed in *E. coli* (tTau assay); recombinant phosphorylated tau expressed in a mammalian system and confirmed by mass spectrometry to display phosphorylation at T181 (pTau181 assay); recombinant GFAP expressed in a mammalian system (GFAP assay); and bovine NfL purified from spinal cord (NfL assay). Due to the lack of international standards, concentrations of calibrators were assigned via biochemical characterization and used to generate a calibration curve for sample quantitation. LLOD was defined as the concentration that provides a signal 2.5 standard deviations above the mean of the blank. LLOQ was defined as the lowest concentration with a CV <20% and a recovery between 80-120%. The samples were codified and randomized so the assay laboratory was blinded to any case information during testing and calculation of concentrations. The samples were distributed over 8 plates per assay, each containing an 8-point calibration curve and QC samples in duplicates and ran over two days. Plasma samples were measured as single replicates using 25 uL of undiluted plasma for the NfL and pTau181 assays, or 25 uL of 5-fold diluted plasma for the GFAP and total tau assays. Reported concentrations of GFAP and total tau were corrected for the 5-fold sample dilution.

### Statistical analysis

Participant demographics and characteristics are presented as mean and standard deviations for continuous variables and frequencies and relative frequencies for categorical variables. Biomarker concentrations were natural log transformed to satisfy assumptions of normal distribution. Values under LLOQ were assigned the lowest quantifiable value of the assay. All reported p-values were adjusted for multiple hypothesis testing using the Benjamini-Hochberg method unless otherwise specified. Differences between diagnostic groups were evaluated using ANOVA adjusting for age, sex, and the biomarker in question followed by Tukey’s Honest Significant Difference as the post-hoc test. Subgroup analyses between different clinical subsets were performed using logistic regression predicting the subgroup in terms of age, sex, and the relevant biomarker. To assess classification utility of the biomarkers, area under the curve (AUC) values were computed using logistic regression models as described above^24^. Effect sizes of each predictor were calculated using Cohen’s d. Correlations between markers and with cognitive scores were assessed with Pearson correlation coefficient or Spearman’s Rho for ordinal data or distributions containing outlier data. To ameliorate the influence of age on biomarkers, levels were residualized in terms of age before correlative analysis of cognitive scores. The above procedures were carried out using the R statistical software version 4.0.4 (R Foundation for Statistical Computing, Vienna, Austria).

### Data availability

Anonymized data not published within this article will be made available by reasonable request from any qualified investigator.

## RESULTS

### Analytical performance of assays

The analytical performance of the four S-PLEX assays is summarized in Table 1. All plasma samples had concentrations exceeding LLOD for all assays. The assay signal was linear with concentration across the full calibration range of the assay. For the three prototype assays (tTau, NfL, and GFAP), 4 QC samples in duplicate with concentrations spanning the assay range were included in each plate. One QC sample in duplicate per plate was used for the commercial pTau181 assay. The reported LLOD, LLOQ, and ULOQ values for GFAP and tTau were adjusted to account for the 5x dilution used with these assays.

**Table 1.** Assay performance.

|  | LLOD<br>(pg/mL) | LLOQ<br>(pg/mL) | ULOQ<br>(pg/mL) | Median conc (Q1-Q3)<br>(pg/mL) | CV for QC<br>samples |
| --- | --- | --- | --- | --- | --- |
| pTau181 | 0.08 | 0.46 | 990 | 1.7 (1.2 – 2.6) | 8% |
| tTau | 0.07 | 0.63 | 2,000 | 10.2 (8.2 – 13.4) | 4-7% |
| NfL | 2.6 | 9.6 | 5,300 | 75 (50 – 116) | 9-13% |
| GFAP | 8.8 | 52 | 10,400 | 170 (119 – 228) | 5-10% |

pTau181=phosphorylated-Tau181; tTau=total Tau; NfL= neurofilament light; GFAP=glial fibrillary acidic protein; LLOD=lower level of detection; L/ULOQ=lower/upper level of quantification; Q1/Q3=Quartile 1/3; CV=coefficient of variation; QC=quality control.

### Diagnostic performance of plasma biomarkers

Diagnostic performance of the four biomarkers under study was evaluated in a high contrast cross-sectional sample (Sample A) including individuals with AD, OND, and CN. Baseline and clinical characteristics are summarized in Table 2A. Initial analysis showed that all four biomarkers (pTau181, tTau, NfL, and GFAP) increased with age in the CN group (eFig 1). There was also an effect of sex for pTau181 in the CN group, with males having higher pTau181 levels than females (p<0.003). This was not observed within the AD or OND groups and was attenuated in the CN group by controlling for age (p<0.02). All subsequent analyses were controlled for age and sex.

**Table 2A.**
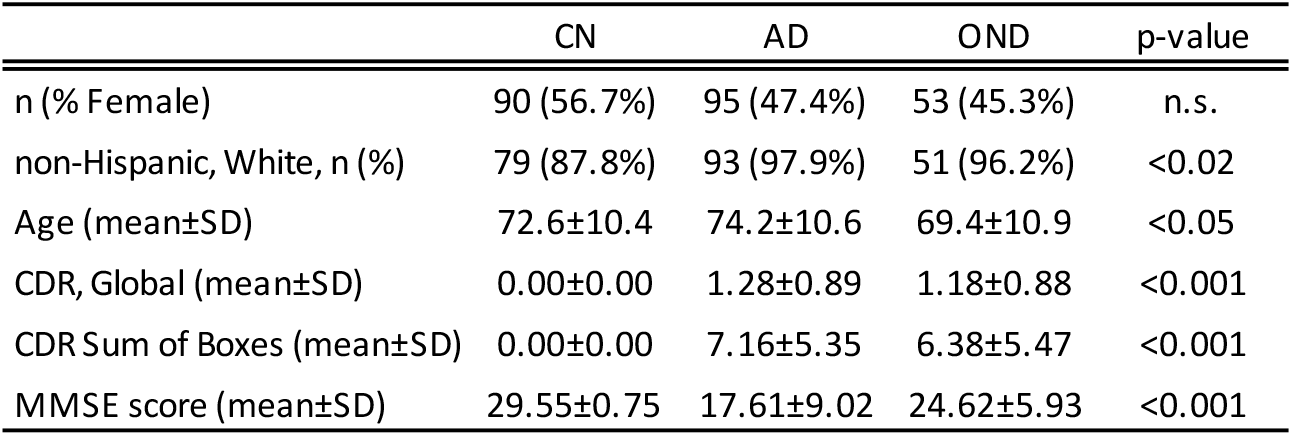
Sample A. “High-Contrast” Diagnostic Sample.

|  | CN | AD | OND | p-value |
| --- | --- | --- | --- | --- |
| n (% Female) | 90 (56.7%) | 95 (47.4%) | 53 (45.3%) | n.s. |
| non-Hispanic, White, n (%) | 79 (87.8%) | 93 (97.9%) | 51 (96.2%) | <0.02 |
| Age (mean±SD) | 72.6±10.4 | 74.2±10.6 | 69.4±10.9 | <0.05 |
| CDR, Global (mean±SD) | 0.00±0.00 | 1.28±0.89 | 1.18±0.88 | <0.001 |
| CDR Sum of Boxes (mean±SD) | 0.00±0.00 | 7.16±5.35 | 6.38±5.47 | <0.001 |
| MMSE score (mean±SD) | 29.55±0.75 | 17.61±9.02 | 24.62±5.93 | <0.001 |

**Table 2B.** Sample B. Longitudinal Prognostic Sample.

|  | MCI_Stable | MCI_Decline | p-value | SCI_Stable | SCI_Decline | p-value | CN_Stable | CN_Decline | p-value |
| --- | --- | --- | --- | --- | --- | --- | --- | --- | --- |
| n (% Female) | 38 (44.7%) | 47 (48.9%) | n.s. | 30 (53.3%) | 19 (63.2%) | n.s. | 39 (58.9%) | 14 (64.3%) | n.s. |
| non-Hispanic, White, n (%) | 35 (92.1) | 45 (95.7) | n.s. | 30 (100.0) | 15 (78.9) | <0.05 | 33 (84.6) | 13 (92.9) | n.s. |
| Age (mean±SD) | 77.8±7.4 | 75.7±8.6 | n.s. | 72.8±5.3 | 79.±6.6 | <0.0001 | 70.8±11.6 | 79.6±6.9 | <0.02 |
| CDR, Global (mean±SD) | 0.50±0.00 | 0.50±0.00 | n.s. | 0.48±0.09 | 0.39±0.21 | <0.05 | 0.00±0.00 | 0.00±0.00 | n.s. |
| CDR Sum of Boxes (mean±SD) | 1.87±0.87 | 2.65±0.89 | <0.001 | 1.15±0.71 | 0.87±0.66 | n.s. | 0.00±0.00 | 0.00±0.00 | n.s. |
| MMSE score (mean±SD) | 28.29±1.43 | 26.12±4.23 | <0.005 | 28.72±1.65 | 27.94±1.64 | n.s. | 29.72±0.46 | 28.29±1.27 | <0.001 |

Abbreviations: CN=cognitive normal controls; AD=Alzheimer's Disease; OND=other neurodegenerative diseases; CDR=Clinical Dementia Rating; MMSE=Mini mental state examination; MCI=mild cognitive decline; SCI=subjective cognitive impairment.

Between group differences in plasma biomarker concentrations demonstrated that participants with AD had roughly 2-fold higher plasma concentrations of pTau181, NfL, and GFAP compared to CN, while tTau concentrations on average were 1.5-fold higher in AD (p<0.001 for all comparisons; Fig 1; Table 3). Analysis of effect sizes showed large standardized mean differences (Cohen’s d: >0.8; Table 3) for all four markers with the largest effect size for pTau181 (Cohen’s d: 1.50). A logistic regression model controlling for age and sex showed an excellent ability for pTau181 to discriminate between AD and CN with an AUC of 0.88 (Fig 2) with 82% sensitivity and 88% specificity at an optimal threshold defined by the Youden index (Table 3). AUCs for differentiating between AD and CN ranged between 0.78-0.83 for tTau, NfL, and GFAP.

**Figure 1.**
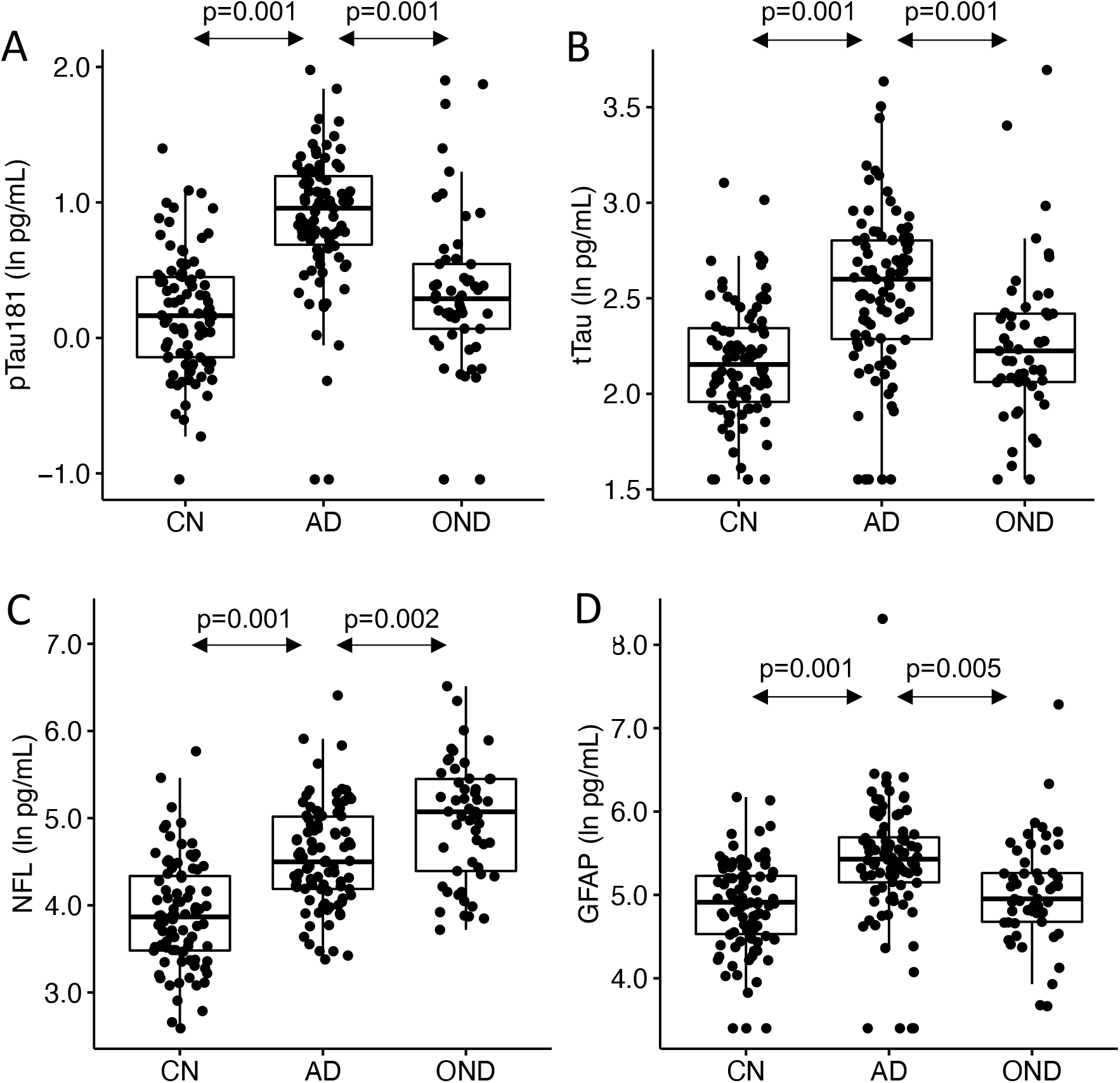
Plasma levels of pTau181 (A), tTau (B), NfL (C), and GFAP (D) in participants with Alzheimer’s disease (AD), other neurodegenerative diseases (OND), and cognitively normal controls (CN). Box plots show median, 25^th^/75^th^ percentile, and smallest/largest value within 1.5x the interquartile below/above the median.

**Table 3.** Clinical performance of the four biomarker assays in Sample A (“High contrast” diagnostic sample)

|  | CN (n=90) |  | AD (n=95) |  | OND (n=53) |  |
| --- | --- | --- | --- | --- | --- | --- |
|  | Mean (SD) | Median (Q1-Q3) | Mean (SD) | Median (Q1-Q3) | Mean (SD) | Median (Q1-Q3) |
| pTau181 (pg/ml) | 1.32±0.63 | 1.18 (0.87-1.57) | 2.67±1.13 | 2.61 (1.97-3.32) | 1.72±1.33 | 1.34 (1.07-1.73) |
| tTau (pg/ml) | 9.2±3.1 | 8.6 (7.0-10.4) | 13.7±5.8 | 13.5 (9.8-16.5) | 10.4±5.9 | 9.3 (7.9-11.2) |
| NfL (pg/ml) | 59.9±46.2 | 47.8 (32.4-76.9) | 127.4±160.0 | 89.9 (65.8-152.0) | 178.2±126.4 | 159.6 (80.9-232.6) |
| GFAP (pg/ml) | 151±82 | 136 (92-187) | 293±413 | 228 (172-299) | 194±201 | 142 (108-193) |

|  | AD vs CN |  |  | AD vs OND |  |  | CN vs OND |  |  |
| --- | --- | --- | --- | --- | --- | --- | --- | --- | --- |
|  | Fold change | Cohen's d | p-value | Fold change | Cohen's d | p-value | Fold change | Cohen's d | p-value |
| pTau181 (pg/ml) | 2.02 | 1.50 | <0.001 | 1.55 | 1.01 | <0.001 | 1.30 | 0.34 | n.s. |
| tTau (pg/ml) | 1.50 | 0.99 | <0.001 | 1.31 | 0.67 | <0.001 | 1.14 | 0.25 | n.s. |
| NfL (pg/ml) | 2.13 | 1.10 | <0.001 | 0.71 | 0.58 | <0.002 | 2.97 | 1.68 | <0.001 |
| GFAP (pg/ml) | 1.94 | 0.83 | <0.001 | 1.51 | 0.54 | <0.005 | 1.29 | 0.28 | n.s. |

|  | Differentiation AD vs CN |  |  | Differentiation AD vs OND |  |  |
| --- | --- | --- | --- | --- | --- | --- |
|  | AUC (95% CI) | %Sensitivity | %Specificity | AUC (95% CI) | %Sensitivity | %Specificity |
| pTau181 | 0.88 (0.83-0.93) | 82% | 88% | 0.78 (0.70-0.87) | 78% | 77% |
| tTau | 0.79 (0.72-0.85) | 66% | 86% | 0.72 (0.63-0.81) | 55% | 83% |
| NfL | 0.83 (0.77-0.89) | 77% | 78% | 0.70 (0.60-0.79) | 87% | 55% |
| GFAP | 0.78 (0.71-0.85) | 74% | 76% | 0.70 (0.61-0.79) | 57% | 81% |

Abbreviations: CN=cognitively normal; AD=Alzheimer's Disease; OND=other neurodegenerative diseases; Q1/Q3=Quartile 1/3; AUC=area under the curve. Reported sensitivity and specificity were determined at the point of maximum sensitivity and specificity determined by the Youden index.

**Figure 2.**
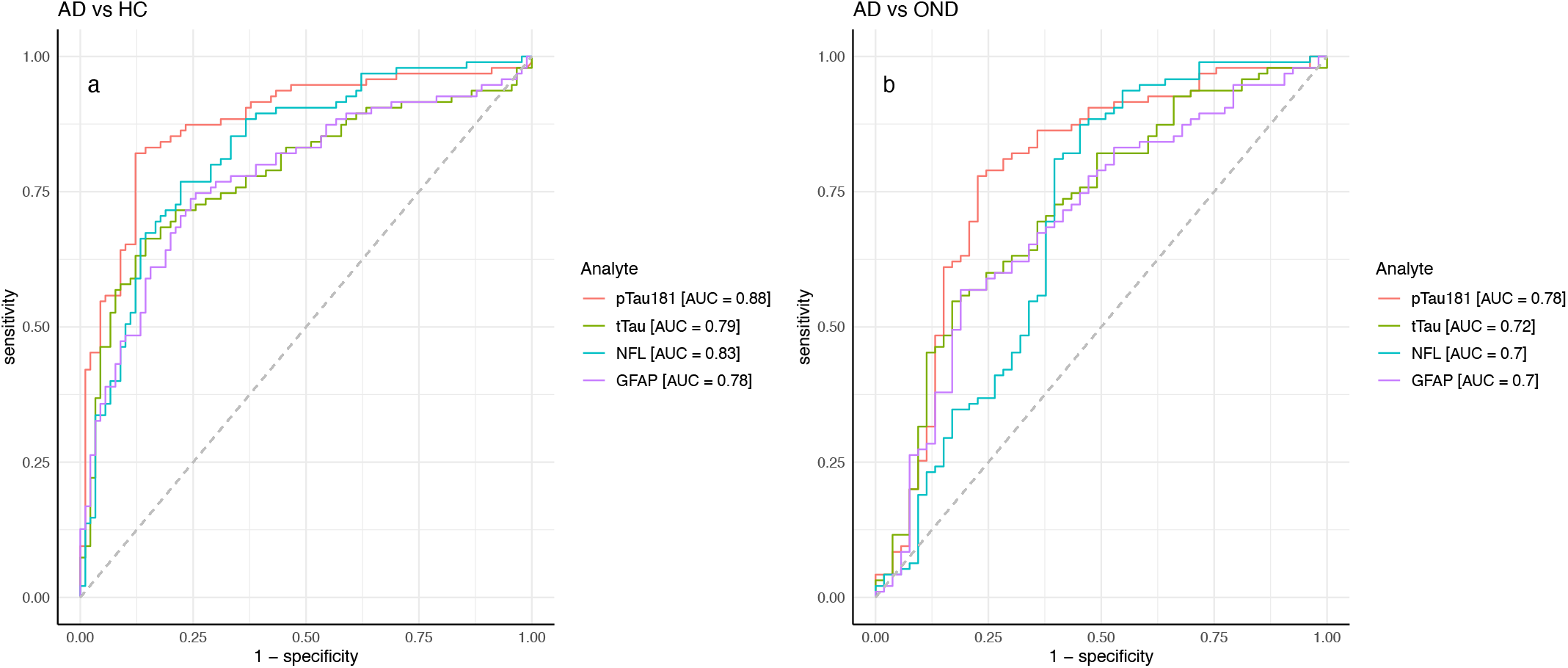
ROC curves for classification of (A) AD vs cognitively normal controls (CN), and (B) AD vs other neurodegenerative diseases (OND), using pTau181, tTau, NfL, and GFAP. Legends show AUC values.

Participants with AD also had roughly 1.3-1.5-fold higher plasma pTau181, tTau, and GFAP concentrations than ONDs (pTau181 and tTau: p<0.001; GFAP: p<0.005; Fig 1; Table 3), again with pTau181 showing the largest fold difference between the groups. A large effect size was observed for pTau181 (Cohen’s d: 1.01; Table 3), while effect sizes were moderate for GFAP (Cohen’s d: 0.54) and tTau (Cohen’s d: 0.67). NfL concentrations were, in contrast, 1.4-fold higher in ONDs compared to AD (p<0.001; Cohen’s d: 1.68), consistent with it being a non-specific marker for neuronal injury^11^. AUCs for differentiating between AD and OND were 0.78 for pTau181 (78% sensitivity and 77% specificity) and 0.70-0.72 for tTau, NfL, and GFAP (Fig 2; Table 3).

### Cross-sectional correlation with disease severity and cognitive function

Next, we assessed if plasma biomarker concentrations were significantly associated with disease severity or global cognitive function at the time of the blood draw in the high contrast diagnostic sample. Clinical dementia severity was assessed by global CDR and CDR sum of boxes (SOB) scores in all participants. Cognitive impairment was evaluated using the Mini-Mental State Examination in 149 participants. A positive association in this analysis was observed for GFAP, which showed moderate correlations with global CDR (Spearman’s rho=0.44; p<0.0001), CDR SOB (rho=0.45; p<0.0001), as well as MMSE (rho=-0.44; p<0.0005). We also observed weak correlations between NfL and CDR SOB (rho=0.22; p<0.05) as well as global CDR (rho=0.19; p<0.06), while no associations were observed for pTau181 or tTau.

### Longitudinal prediction of cognitive decline

We used Sample B to investigate whether plasma biomarker levels at baseline can predict clinical progression during the next four years of follow up in older adults with no cognitive impairment (CN), SCI, or MCI with a consensus clinical diagnosis of AD as the likely underlying etiology. Baseline characteristics are summarized in Table 2B and the cognitive trajectories of the different groups are illustrated in eFig 2. There were no differences in age or sex between MCI-stable and MCI-decline participants, while the CN- and SCI-decline groups tended to be older than their stable counterparts (Table 2B). Age and sex were therefore included as covariates in the analysis.

Participants with MCI who progressed to AD dementia (MCI-decline) had higher baseline plasma concentrations of pTau181, NfL, and GFAP compared to MCI-stable participants with the largest fold change for pTau181 and GFAP (1.7 and 1.6-fold increase, respectively; p<0.0005 for both comparisons; Fig 3; Table 4). NfL levels were significantly higher in MCI-decline (p<0.05) compared to stable participants but the difference was modest (1.1-fold increase). This was reflected by a large effect size for pTau181 (Cohen’s d: 1.15) and GFAP (Cohen’s d: 1.03), while the effect size for NfL was considerably smaller (Cohen’s d: 0.32). There were no differences in pTau181 or GFAP levels between MCI-decline [pTau181 (mean±SD): 2.88±1.44 pg/mL; GFAP: 213±92 pg/mL] and the group of AD participants from the high contrast sample (Sample A; pTau181: 2.67±1.13 pg/mL; p=0.65; GFAP: 292±413 pg/mL; p=0.49), while MCI-stable participants had significantly lower pTau181 and GFAP levels compared to both the other two groups (pTau181: 1.73±1.13 pg/mL; GFAP: 137±68 pg/mL; p<0.001 for all comparisons). pTau181 and GFAP showed the best ability to discriminate between MCI-decline and MCI-stable with an AUC for pTau181 of 0.82 with 85% sensitivity and 79% specificity and an AUC for GFAP of 0.81 with 81% sensitivity and 66% specificity (Table 4).

**Figure 3.**
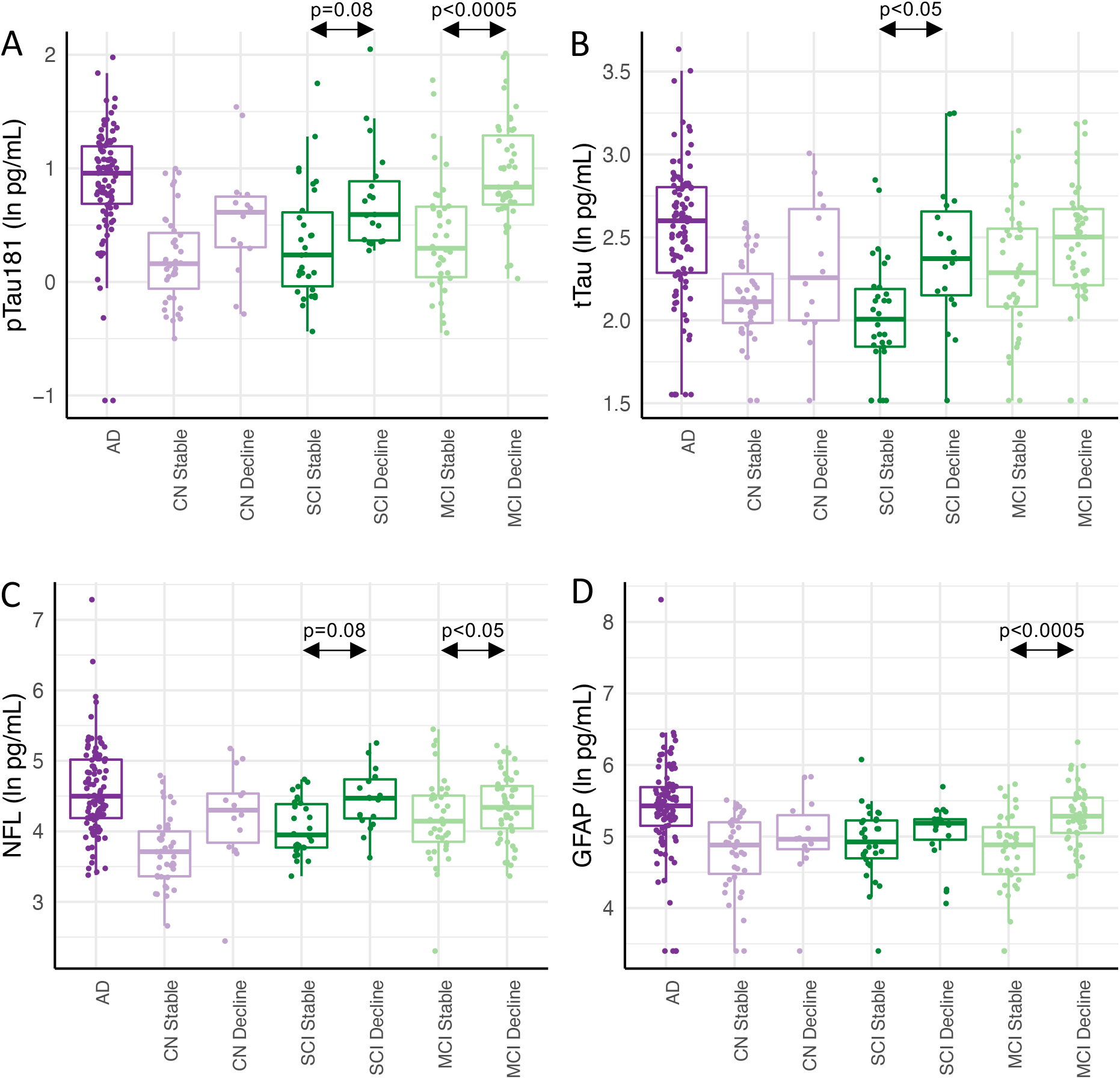
Plasma levels of pTau181 (A), tTau (B), NfL (C), and GFAP (D) in participants with mild cognitive impairment (MCI), subjective cognitive impairment (SCI), or no cognitive impairment (CN) stratified by the presence (decline) or absence (stable) of cognitive decline during 4 years of follow-up. Biomarker levels in all participants with autopsy- or biomarker confirmed AD are included as a reference. Box plots show median, 25^th^/75^th^ percentile, and smallest/largest value within 1.5x the interquartile below/above the median.

**Table 4.** Clinical performance of the four biomarker assays in Sample B (Longitudinal prognostic sample)

|  | MCI Decline (n=47) |  | MCI Stable (n=38) |  | Fold change | Cohen's d | p-value |
| --- | --- | --- | --- | --- | --- | --- | --- |
|  | Mean (SD) | Median (Q1-Q3) | Mean (SD) | Median (Q1-Q3) |  |  |  |
| pTau181 (pg/ml) | 2.88 (1.44) | 2.31 (1.94-3.80) | 1.73 (1.13) | 1.35 (1.03-1.95) | 1.67 | 1.15 | <0.0005 |
| tTau (pg/ml) | 12.5 (4.6) | 12.2 (9.1-14.5) | 10.8 (4.3) | 9.8 (8.0-12.9) | 1.15 | 0.39 | n.s. |
| NfL (pg/ml) | 85 (38.4) | 76.7 (55-106.6) | 76.7 (47.8) | 63 (46.3-91.4) | 1.11 | 0.32 | <0.05 |
| GFAP (pg/ml) | 213 (93) | 197 (155-257) | 137 (69) | 132 (87-174) | 1.56 | 1.03 | <0.0005 |

|  | SCI Decline (n=14) |  | SCI Stable (n=39) |  | Fold change | Cohen's d | p-value |
| --- | --- | --- | --- | --- | --- | --- | --- |
|  | Mean (SD) | Median (Q1-Q3) | Mean (SD) | Median (Q1-Q3) |  |  |  |
| pTau181 (pg/ml) | 2.38 (1.53) | 1.81 (1.43-2.54) | 1.62 (1.05) | 1.27 (0.93-1.88) | 1.46 | 0.82 | 0.08 |
| tTau (pg/ml) | 12.1 (5.7) | 10.7 (8.4-14.8) | 8.1 (3.1) | 7.4 (6.3-9.0) | 1.51 | 1 | <0.05 |
| NfL (pg/ml) | 92.8 (39.7) | 87.4 (63.8-117.4) | 61.5 (24.3) | 51.9 (43.3-81.3) | 1.51 | 1.02 | 0.08 |
| GFAP (pg/ml) | 166 (57) | 179 (134-189) | 152 (75) | 138 (108-186) | 1.10 | 0.28 | n.s. |

|  | CN Decline (n=14) |  | CN Stable (n=39) |  | Fold change | Cohen's d | p-value |
| --- | --- | --- | --- | --- | --- | --- | --- |
|  | Mean (SD) | Median (Q1-Q3) | Mean (SD) | Median (Q1-Q3) |  |  |  |
| pTau181 (pg/ml) | 1.99 (1.16) | 1.85 (1.34-2.16) | 1.33 (0.57) | 1.17 (0.87-1.57) | 1.50 | n.d. | n.s. |
| tTau (pg/ml) | 11.0 (4.7) | 9.6 (7.3-14.7) | 8.7 (2.1) | 8.3 (7.2-10.2) | 1.27 | n.d. | n.s. |
| NfL (pg/ml) | 82.1 (47.3) | 73.9 (43.7-94.1) | 47.1 (25.3) | 40.9 (28.7-56.9) | 1.74 | n.d. | n.s. |
| GFAP (pg/ml) | 168 (88) | 143 (123-212) | 134 (59) | 132 (84-181) | 1.25 | n.d. | n.s. |

| <u>Differentiation MCI Stable vs Decline</u> |  |  |  |
| --- | --- | --- | --- |
|  | AUC (95% CI) | %Sensitivity | %Specificity |
| pTau181 | 0.82 (0.72-0.92) | 85% | 79% |
| tTau | 0.63 (0.51-0.75) | 34% | 95% |
| NfL | 0.68 (0.56-0.79) | 74% | 61% |
| GFAP | 0.81 (0.72-0.90) | 81% | 66% |

Abbreviations: MCI=mild cognitive impairment; SCI=subjective cognitive impairment; CN=cognitively normal; Q1/Q3=quartile 1/3; AUC=area under the curve. Reported sensitivity and specificity were determined at the point of maximum sensitivity and specificity determined by the Youden index.

Individuals with SCI frequently fluctuate in their yearly cognitive status^25^ which led us to only consider participants with SCI as SCI-decliners if they had at least two consecutive follow up visits in which their consensus diagnosis was MCI and their global CDR was 0.5, without subsequent return to SCI status. Using these criteria, we identified 19 SCI-decline and 30 SCI-stable participants. Levels of pTau181, tTau, and NfL were all increased (Fig 3; Table 4) with 1.5-fold higher levels in SCI-decline compared to SCI-stable participants, but the only differences which remained significant after multiple testing correction was tTau (p<0.05; Cohen’s d:1.00) presumably due to the relatively small sample size and unreliable diagnostic categories. There were no statistically significant differences in baseline biomarker concentrations between CN individuals with and without cognitive decline, but the number of CN-decline participants was also small (n=14; Table 4).

### Correlations among AD biomarkers

We observed strong correlations between levels of pTau181 and tTau not only among individuals with AD (rho=0.54, p<0.0001), but also among CN and ONDs (rho=0.55 and 0.58, respectively; p<0.0001 for both comparisons). pTau181 also correlated moderately with GFAP (rho=0.35, p<0.001) and NfL (rho=0.35, p<0.001) within the AD group, but this correlation was lost among ONDs (GFAP: rho=0.08, p=0.3; NfL: rho=0.06, p=0.3) likely reflecting that disease mechanisms other than amyloid and tau pathology also increase GFAP and NfL levels in these individuals. tTau also correlated with GFAP (rho=0.30, p<0.005) and NfL (r=0.54, p<0.0001) within the AD group, but not among the ONDs (GFAP: rho=0.10, p=0.5; NfL: rho=0.32, p=0.5) similar to what was observed for pTau181.

### Classification of Evidence

This study provides Class II evidence that plasma levels of pTau181, tTau, NfL, and GFAP are associated with AD and that pTau181 and GFAP are associated with progression from MCI to AD dementia.

## DISCUSSION

We describe the diagnostic and prognostic value of four plasma biomarkers of AD neuropathology (pTau181, tTau, NfL, and GFAP) in 366 participants in the longitudinal cohort of the Massachusetts ADRC. These cases spanned normal aging, subjective cognitive impairment, and mild cognitive impairment with and without progression, dementia due to AD, and other neurodegenerative diseases. We confirm previous findings that all four biomarkers provide predictive diagnostic value for AD, and we extend emerging findings that pTau181, GFAP and to a lesser degree NfL inform prognosis with their higher levels predicting decline in participants with MCI.

pTau181, tTau, NfL, and GFAP were measured in plasma using novel ultrasensitive ECL-based MSD immunoassays. The assays performed well and detected higher plasma levels of all four biomarkers in individuals with AD compared to both individuals with normal cognition and, with the exception of NfL, individuals with other non-AD neurodegenerative diseases. The best performance was observed for pTau181, which could discriminate between AD and CN with an AUC of 0.88 and between AD and non-AD neurodegenerative diseases with an AUC of 0.78. This diagnostic accuracy between AD and CN is comparable to that originally observed using SIMOA assays on the Quanterix platform and what subsequently has been reported in several studies^6, 8, 26, 27^ but nominally lower than what was recently described when comparing the commercial SIMOA pTau181 assay with two novel prototype pTau181 SIMOA assays developed by Eli Lilly and ADx NeuroSciences^5^. These relatively small discrepancies in diagnostic accuracy may be due to different patient populations, the prevalence of tau pathology, and the diagnostic criteria used to define AD. It is also possible that our sample included some misclassified healthy controls with asymptomatic AD pathology as we only required biological verification of the AD and non-AD neurodegenerative groups in the diagnostic “high contrast” group.

We also observed good performance of the other three biomarkers under study with a diagnostic accuracy between AD and CN in the 0.78-0.83 range. The sensitivity of ECL assays has not previously been adequate to quantify plasma NfL levels in AD^28^, but advances in ultrasensitive detection in the ECL assays have made it possible to detect levels in the single picogram range using the current enhanced assay, which has an LLOQ of 9.6 pg/mL. All participants in the current study had measurable NfL plasma levels. Notably, while the vast majority of other assays use the “gold standard” antibody pair developed by Uman Diagnostics [anti NF-L mAb 47:3 (UD1) and anti NF-L mAb 2:1 (UD2)]^29^, the current assay uses a novel antibody pair developed by MSD. With this new assay, we confirm the observations that average plasma NfL levels are higher in AD compared to CN^30^, but that other neurodegenerative disorders have equally high or higher plasma NfL level^31^. We noted marginally higher NfL levels at baseline in individuals with MCI who progressed to dementia during follow up, but the prognostic accuracy was low, consistent with previous studies that have failed to show elevated baseline NfL levels in individuals with MCI who progress to AD dementia^32, 33^.

As a biofluid biomarker, CSF tTau is believed to reflect amyloid-β-induced tau secretion from neurons in AD^34^. CSF tTau levels are also temporarily increased after traumatic brain injury^35^ and highly increased in diseases with rapid and extensive neurodegeneration such as Creuzfeldt-Jakob disease^36^, where increased tau levels are thought to indicate release of tau with neuronal death, reflecting a different pathological process than in AD^34^. Increased plasma levels of tTau have previously been described in AD, but differences in average levels between AD and control groups have been small, their distributions largely overlapping, and plasma tTau levels were only weakly correlated with CSF tTau limiting the usefulness of plasma tTau as measured in that assay as a diagnostic marker in AD^37-39^. The novel ECL assay tested here confirmed that individuals with AD have increased tTau levels in plasma compared to CN but showed better separation between the groups than previous studies, with 1.5-fold higher average concentrations in AD compared to CN and a diagnostic accuracy of 0.79. It is thought that the discrepancy in tTau levels between CSF and plasma may be explained by proteolytic degradation of tau in the blood or by contribution of peripheral tau^40^. We observed a strong correlation between tTau and pTau181 levels using the ECL assays in our study suggesting that the observed tTau levels do, at least in part, reflect AD pathology. It can be speculated that the epitopes detected by the MSD antibodies detect tau fragments that are more resistant to protease degradation or that are more CNS specific than the previous assays in analogy to the recently described N-terminal tau fragment NT-1 which was highly predictive of future cognitive decline and pathological tau accumulation in clinically normal elderly^41^.

GFAP was the only biomarker in our study that was significantly associated with clinical severity and cognitive status in cross-sectional analysis. We also noted higher plasma levels of GFAP at baseline in individuals with MCI who progressed to dementia during follow up compared to stable MCI with an AUC between the two groups of 0.81. Plasma GFAP levels have recently been shown to be associated with progression from MCI to AD dementia^42^ and were increased in cognitively unimpaired adults at risk of AD^15, 16, 43^. Plasma GFAP has furthermore been shown to correlate with disease severity and neuropsychological test performance^15, 16, 43, 44^. Increased GFAP expression is detected in astrocytes around amyloid-β plaques in AD brains^45^ and GFAP has been proposed as an early and specific marker for amyloid-β induced astrocyte pathology in AD due to its association with amyloid-β PET signal not only in individuals with MCI or dementia but also in cognitively unimpaired individuals, while no association was observed with tau PET burden in the absence of amyloid-β pathology^15,16^. It was suggested that GFAP levels in plasma reflect amyloid-β pathology better than CSF levels as only plasma GFAP could differentiate between amyloid-β positive and negative CN individuals^15^. However, it is also clear that increased plasma GFAP may not only be present in AD, as astrocytosis occurs in response to many different pathological processes in the CNS^46^, and increased plasma GFAP levels have also been linked with conditions such as traumatic brain injury, stroke, FTD, and multiple sclerosis^44, 47-49^. It remains to be determined how plasma GFAP can be utilized in AD, but it can be speculated that its early and gradual increase together with its association with amyloid-β pathology would make it useful for predicting and following disease progression both in clinical practice and during clinical trials, as well as to support inclusion of individuals with early AD^50^.

The strengths of this study include a well characterized diagnostic sample with autopsy- or biomarker verified diagnosis of the participants with AD and non-AD neurodegenerative diseases, and the careful selection of individuals with at least four years of follow up for the prognostic analyses. Adequate sample sizes were used in the main groups being compared, which were similar in age and other demographic attributes. Initial analysis indicated an effect of age on biomarker levels and all analysis was therefore controlled for age. Limitations include the small numbers of decliners in the CN and SCI groups and the frequent fluctuations in their clinical diagnoses, rendering some uncertainty in findings in these groups and we include them with this caveat. Some of the participants in the longitudinal prognostic sample may have been misclassified due to the lack of CSF and PET imaging biomarkers of amyloid-ß and tau. It is also possible that comorbid pathologies (i.e., vascular, Lewy body, TDP-43) may contribute to the progression in this sample but could not be accounted for due to the lack of autopsy confirmation or relevant biomarkers. Lastly, our study population consisted largely of white non-Hispanics, which limits the generalizability of the results.

The new generation of ultrasensitive ECL assays measuring plasma AD biomarkers evaluated in this study provided sufficient accuracy to serve both as diagnostic and prognostic biomarkers in AD and can be measured using technology currently widely available in research laboratories. The rapid development of ultrasensitive assays for measuring AD biomarkers in blood holds promise to transform clinical practice and clinical research in providing affordable and easily accessible assays to assist in diagnosis and prognosis that can be implemented not only in large, centralized settings but equally well in community settings and smaller laboratories lacking the resources to procure expensive specialized equipment. With further investigation and development, they may further open the possibility for screening of asymptomatic individuals for preventative interventions.

## ACKNOWLEDGEMENTS

We thank all study participants and their families for their invaluable contributions. The Harvard Biomarkers Study (“HBS”); https://www.bwhparkinsoncenter.org) is a collaborative initiative of Brigham and Women’s Hospital and Massachusetts General Hospital, co-directed by Dr. Clemens Scherzer and Dr. Bradley T. Hyman. The HBS Study Investigators are: Harvard Biomarkers Study Biobank: Co-Directors: Brigham and Women’s Hospital: Clemens R. Scherzer, Massachusetts General Hospital: Bradley T. Hyman; Investigators and Study Coordinators: Brigham and Women’s Hospital: Idil Tuncali, Elena Abatzis, Michael T. Hayes, Aleksandar Videnovic, Nutan Sharma, Vikram Khurana, Claudio Melo De Gusmao, Reisa Sperling; Massachusetts General Hospital: John H. Growdon, Michael A. Schwarzschild, Albert Y. Hung, Alice W. Flaherty, Deborah Blacker, Anne-Marie Wills, Steven E. Arnold, Ann L. Hunt, Nicte I. Mejia, Anand Viswanathan, Stephen N. Gomperts, Mark W. Albers, Maria Allora-Palli, David Hsu, Alexandra Kimball, Scott McGinnis, John Becker, Randy Buckner, Thomas Byrne, Maura Copeland, Bradford Dickerson, Matthew Frosch, Theresa Gomez-Isla, Steven Greenberg, Julius Hedden, Elizabeth Hedley-Whyte, Keith Johnson, Raymond Kelleher, Aaron Koenig, Maria Marquis-Sayagues, Gad Marshall, Sergi Martinez-Ramirez, Donald McLaren, Olivia Okereke, Elena Ratti, Christopher William, Koene Van Dij, Shuko Takeda, Anat Stemmer-Rachaminov, Jessica Kloppenburg, Catherine Munro, Rachel Schmid, Sarah Wigman, Sara Wlodarcsyk; Data Coordination: Brigham and Women’s Hospital: Thomas Yi; Biobank Management Staff: Brigham and Women’s Hospital: Grace Greco. The content is solely the responsibility of the authors and does not necessarily represent the official views of supporting organizations.

---

**eFigure 1.**
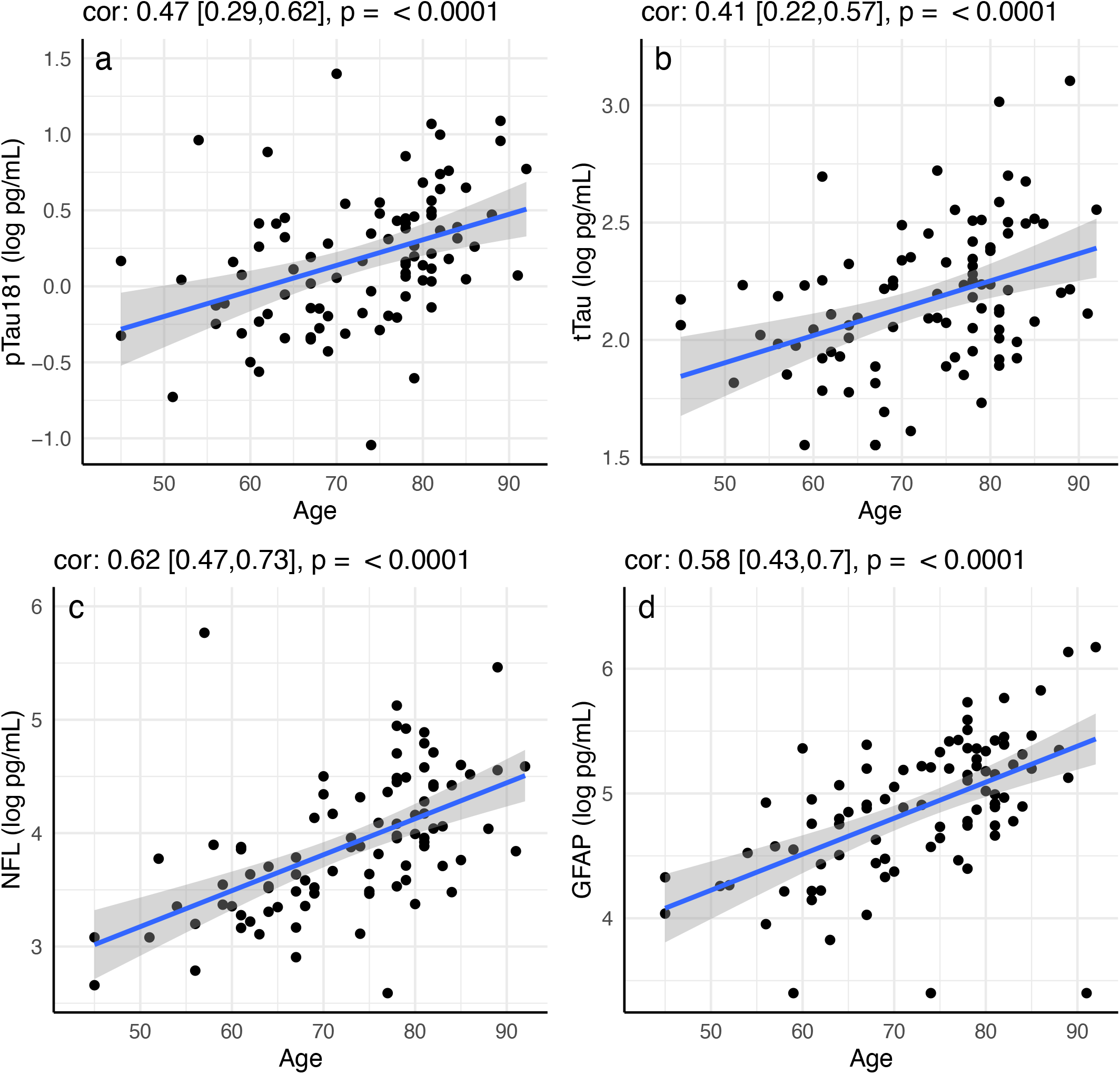
Correlation between age and plasma levels of (A) pTau181, (B) tTau, (C) NfL, and (D) GFAP using Spearman rank-order correlation. Spearman’s rho with 95% confidence interval and p-values are show on top of each panel.

**eFigure 2.**
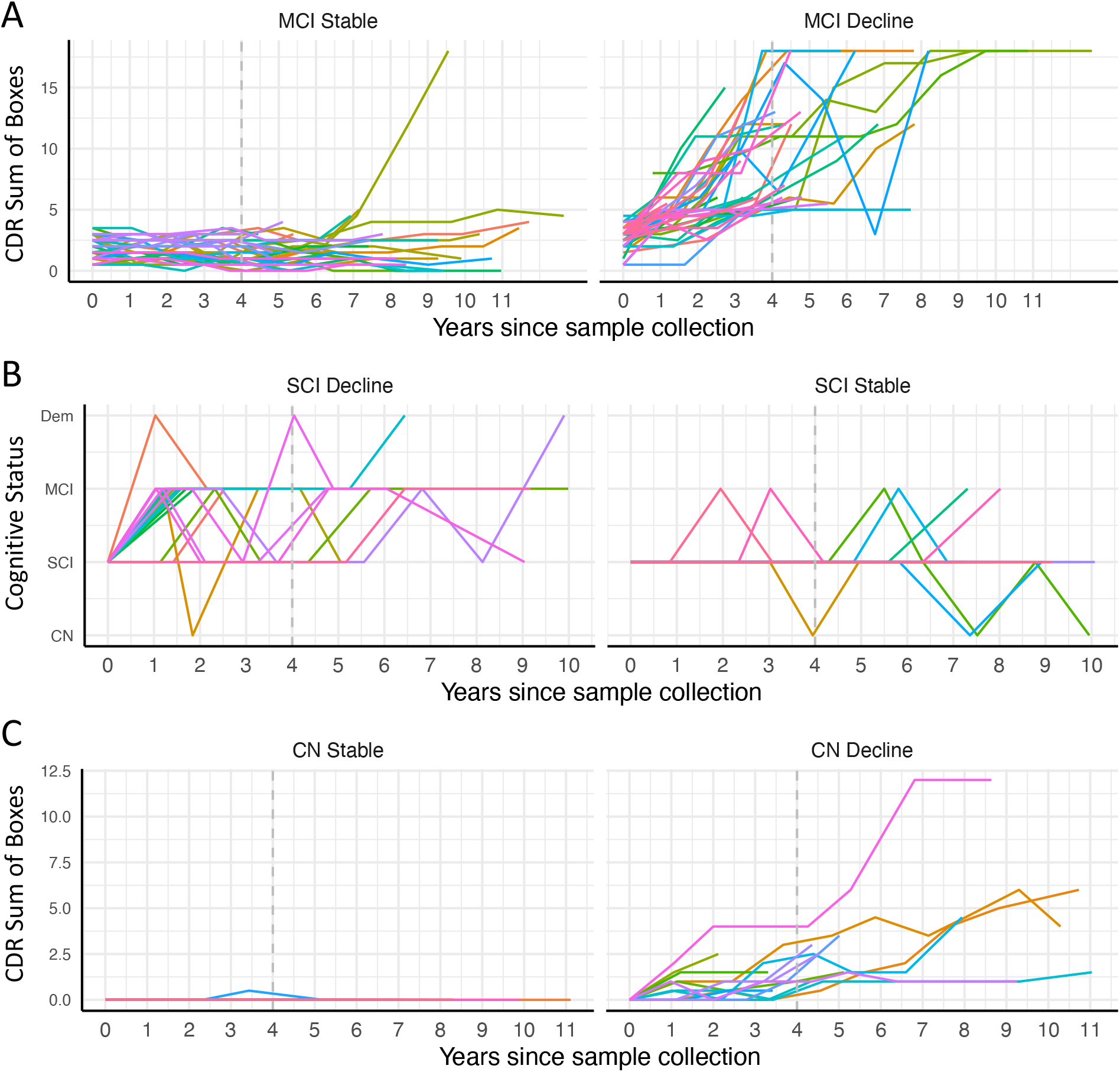
Trajectory of CDR Sum of Boxes scores or Cognitive Status over longitudinal visits in participants with (A) mild cognitive impairment (MCI), (B) subjective cognitive impairment (SCI), or (C) no cognitive impairment (CN) at the time of blood draw classified as stable or decliners based on their cognitive trajectories during 4 years of follow-up (indicated with dashed gray line).

